# Factors Associated with Good Patient Outcomes Following Convalescent Plasma in COVID-19: A Prospective Phase II Clinical Trial

**DOI:** 10.1101/2020.08.27.20183293

**Authors:** Danyal Ibrahim, Latha Dulipsingh, Lisa Zapatka, Reginald Eadie, Rebecca Crowell, Kendra Williams, Dorothy B. Wakefield, Lisa Cook, Jennifer Puff, Syed A. Hussain

**Affiliations:** Trinity Health Of New England, Hartford, Connecticut, USA; Diabetes Center, Saint Francis Hospital, Hartford, Connecticut, USA; Trinity Health Of New England, Hartford; Research Department, Saint Francis Hospital, Hartford; Diabetes Center, Saint Francis Hospital, Hartford

**Author notes:** **Corresponding author** Danyal Ibrahim, M.D., M.P.H. Trinity Health Of New England, 114 Woodland Street, Hartford, CT 06105.

**Keywords:** Adult, Humans, Length of Stay, Patient Discharge, Hospital Mortality, Respiratory Distress Syndrome, Adult, Prospective Studies, COVID-19, severe acute respiratory syndrome coronavirus 2, Hospitals, Respiration, Ventilation, Plasma, Disease Progression, Immunoglobulin G, Plasma, COVID-19 serotherapy, Clinical Trial, Phase II

## Abstract

We conducted a prospective single-arm open-label phase II clinical trial assessing the safety and efficacy of convalescent plasma in hospitalized COVID-19 patients. Convalescent plasma with sufficient IgG titer (1:320) obtained from recovered donors was administered to adult patients with either severe or critical COVID-19 illness. Primary outcomes were adverse events in association with plasma administration, and hospital mortality. Secondary outcomes included disease progression, recovery, length of stay, and hospital discharge. Of the 38 patients included in the analysis, 24 (63%) recovered and were discharged, and 14 (37%) died. Patients who received convalescent plasma early in the disease course (severe illness group) as compared to the patients that received convalescent plasma later in disease progression (critical illness group) had significantly lower hospital mortality 13% vs 55% (p<0.02) and shorter mean hospital length of stay 15.4 vs 33 days (p<0.01). One patient experienced a transient transfusion reaction. No other adverse effects of convalescent plasma infusion were observed. Our results suggest that convalescent plasma is safe and has the potential for positive impact on clinical outcomes including recovery and survival if given to patients early in the course of COVID-19 disease.

## Introduction

Coronavirus Disease 2019 (COVID-19) is a viral respiratory syndrome caused by severe acute respiratory syndrome coronavirus 2 (SARS-CoV-2). As of July 24, 2020, this novel virus was discovery in Wuhan City, Hubei Province, China, in December 2019, confirmed cases have risen to more than 14,000,000 worldwide and more than 600,000 people have died (*1*). Currently, no cure or standard treatment for COVID-19 exists.

The majority of people with COVID-19 experience an asymptomatic, mild, or manageable course of disease (*2, 3*). The most common symptoms include fever, cough, fatigue, dyspnea, headache, diarrhea, fatigue, myalgia, and/or loss of taste and smell (*4, 5*). However, 19% of those who are infected with the virus become severely or critically ill (*2*). Life threatening illness occurs when the virus triggers a progressive hyper-immune response or “cytokine storm” progressing to acute respiratory distress syndrome (ARDS), cardiac injury, thrombotic complications, septic shock and/or organ failure (*6-9*). Estimated mortality among patients admitted to the intensive care unit (ICU) with severe or critical illness ranges from 34.8% to 41.6% (*10, 11*). Risk of death and disease severity increase with older age, obesity and chronic disease such as hypertension, diabetes and cardiovascular disease (*7-9, 12-14*).

In March of 2020, The US Food and Drug Administration (FDA) solicited investigational new drug applications to test the safety and efficacy of convalescent plasma therapy for patients with severe or life-threatening COVID-19 (*15*). Convalescent plasma is derived from the blood of recovered patients and is a rich source of antibodies. When administered to patients who are ill with the same disease, the plasma may aid recovery by conferring passive immunity and neutralizing the pathogen (*15*). The therapy showed promise during outbreaks of other novel viral respiratory syndromes, including two caused specifically by coronavirus (SARS-CoV in 2003 and Middle Eastern Respiratory Syndrome (MERS) in 2012) (*16, 17*). Data showed that convalescent plasma might be most effective when given earlier in the course of disease, but research was limited to small observational studies and much remains unknown (*16, 17*).

Preliminary data from clinical trials and observational studies targeting COVID-19 suggest that administration of convalescent plasma may reduce mortality, hospital length of stay, and time on mechanical ventilation with minimal adverse side-effects in patients with severe or life-threatening disease (*18-23*). Consistent with earlier studies, treatment may be most efficacious for severe COVID-19 when administered closer to symptom onset (*21-25*). The purpose of this study is to describe the course of illness among 38 patients hospitalized with severe or life-threatening COVID-19 who received convalescent plasma as part of an FDA-approved Phase 2 clinical trial. Specifically, the study will assess their hospital course in the context of demographics, disease onset, symptomology, illness severity, and disease progression.

## Material and Methods

This study is an FDA-approved prospective single-arm open-label Phase II clinical trial (NCT04343261) assessing the safety and efficacy of convalescent plasma (IND #19805) on the clinical course of adult patients hospitalized with severe COVID-19. https://clinicaltrials.gov/ct2/show/NCT04343261?term=plasma&cond=COVID-19&cntry=US&draw=3&rank=19

### Patients

Subjects were recruited from four regional hospitals in Connecticut and Massachusetts between period of April 20, 2020 and June 8, 2020. Patients were considered eligible for the study if they were between the ages of 18 and 90, hospitalized, severely or critically ill with confirmed COVID-19 through nasopharyngeal swab real time PCR (RT-PCR). Illness severity was defined as follows: Mild COVID-19 was defined as symptoms with no clinical signs of moderate, severe, or critical disease; moderate illness was defined as respiratory rate ≥ 20 breaths per minute and oxygen saturation > 93%; severe illness was defined as any of the following: respiratory frequency ≥ 30/minute, blood oxygen saturation ≤ 93%, partial pressure of arterial oxygen to fraction of inspired oxygen ratio < 300, and lung infiltrates > 50% within 24 to 48 hours; evidence of critical illness included respiratory failure, septic shock, or multi-organ dysfunction or failure (*15, 26*).

Subjects who met eligibility criteria were referred by their treating physicians. Patients were enrolled regardless of previous treatment or therapies for COVID-19, including experimental medications and therapies administered off-label. Informed consent was provided by either the patient or the patient’s legally authorized representative (LAR). Once a patient or the patient’s LAR provided informed consent and the patient’s ABO blood type was determined, compatible convalescent plasma was administered in 2 consecutive 200 mL infusions. Each unit was transfused for the duration of 1 hour, 1 to 2 hours apart. If the patient received plasma with undetectable antibodies, the patient was re-dosed with a unit of plasma with adequate antibody titer (1:320). Recipients were monitored and all adverse reactions or events were recorded whether or not they were related to the plasma infusion. The protocol was approved by the Trinity Health Of New England Institutional Review Board (#SFH-20-23).

### Convalescent Plasma

Convalescent plasma was obtained from adult donors who were confirmed positive and had recovered from SARS-CoV-2. All donors screened negative for the virus using a nasal swab (RT-PCR) and had IgG titers >6.5 arbitrary units per mL (AU/mL; equivalent to 1:320). Plasma was collected by apheresis at an established blood donation center following standard operating procedures and 21 CFR 630.10 requirements. Plasma was frozen within 24 hours of collection and labeled for investigational use and ABO typing.

### Data and Data Sources

Demographic, clinical and outcomes data were prospectively collected from electronic patient medical records at each of the four hospitals. Descriptive data included sex, age, race, ethnicity, smoking status, functional status, comorbidities, living situation, and means of arrival to the hospital. Initial presentation to the Emergency Department included self-reported symptoms, vital signs, degree of respiratory distress, and need for oxygen supplementation and resuscitation. Initial chest X-ray findings, and laboratory markers of sepsis, inflammatory response, immune deficiency and organ dysfunction were recorded. The clinical course during hospital stay was prospectively captured by tracking changes in oxygenation (FiO2), need for invasive ventilation, ICU level care and types of essential medications given. Patient clinical status progression and recovery were prospectively monitored by capturing days on invasive ventilation, intubation, extubation, discharge alive, and death during hospitalization.

### Outcomes and Data Analysis

Primary clinical outcomes were rate of adverse events associated with convalescent plasma administration, and hospital mortality. Secondary outcomes included disease progression, recovery, length of hospital stay, and hospital discharge. Primary and secondary clinical outcomes were compared between 2 groups based on severity of illness (*15, 26*) at the time of plasma infusion: 1) patients with severe illness, who had not progressed to ARDS at the time of enrollment, and 2) patients whose condition had progressed to critical illness at the time of enrollment. Patients were excluded from the analysis if they were transferred to another acute hospital; did not receive convalescent plasma with adequate antibody titer; or care was withdrawn and patient received comfort care only within 5 days of plasma administration.

### Statistical Analysis

Descriptive statistics included means, medians and proportions as appropriate based on variable, sample size and distribution. Descriptive variables included demographic characteristics, clinical parameters, and time from illness onset and hospitalization to plasma transfusion. Due to the small sample size, both parametric and nonparametric statistics were used in the analysis as appropriate. Continuous variables were compared using t-tests, and categorical variables using Chi-square analyses and Fisher’s exact test when cell-sizes were small. SAS software version 9.4 (SAS Institute Inc., Cary, NC, USA) was used for analyses. Outcomes were considered statistically significant at p< 0.05.

## Results

### Plasma Recipients

A total of 46 patients (Figure 1) with RT-PCR-confirmed COVID-19 were enrolled in the study. Eight 8/46 (17%) patients were excluded from the analysis for the following reasons: 2 received convalescent plasma with no detectable antibody titer; 1 was transferred to another hospital; 5 were made comfort care only (CMO) and medical care was withdrawn within 5 days of plasma administration. The remaining patients (n=38) included in this analysis received convalescent plasma with adequate antibody titer of 1:320 (30 received 2 units, 5 were re-dosed with 1 unit, 1 received 1 unit).

**Figure 1.**
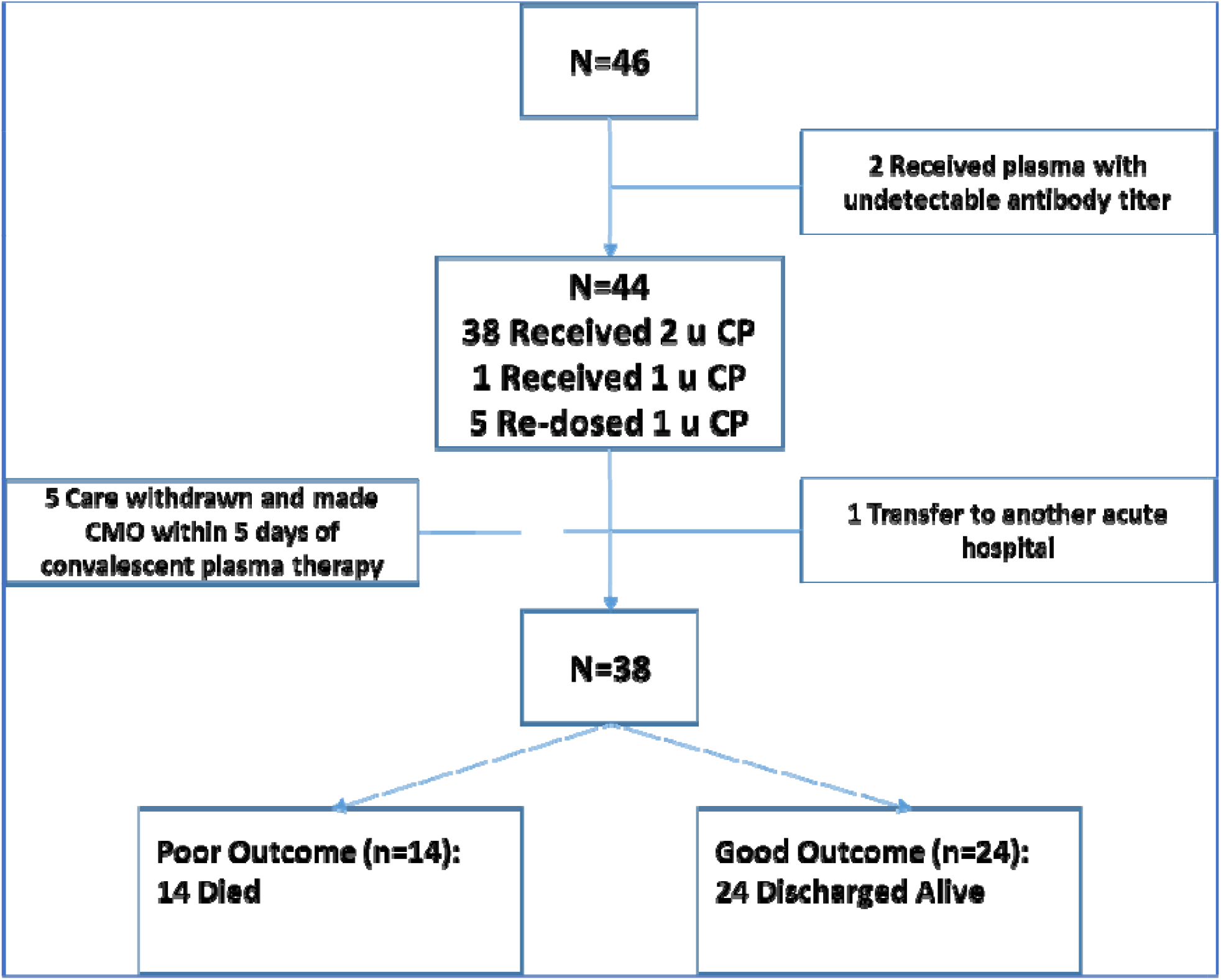
Flowchart of Patients and Outcomes

Patient demographics, clinical presentation, hospital course, and clinical outcomes are shown in tables 1 through 5. Mean age was 63 years (95% CI 59–70), 53% were males, 34% black, 32% white, and 34% were Hispanic; 56% of the patients were from Connecticut, 37% from Harford County, 19% New Haven County, and 42% were from Hampden County, Massachusetts (Table 1). More than 68% had been diagnosed with hypertension and nearly half (47.4%) with diabetes mellitus; overall, 31.5% had three or more comorbidities (Table 2). As shown in Table 3, mean days from onset of symptoms to hospitalization was 7.3 days (95% CI 6.4-8.2), with the most common symptoms at admission being fever, cough and dyspnea. With the exception of one patient who arrived in critical condition, subjects presented initially to the hospital with moderate to severe COVID-19 pneumonia without evidence of ARDS or requiring invasive ventilation support at the time of admission (Table 4). The most common laboratory abnormalities on admission included severe rise in inflammatory markers (C-reactive protein (CRP) ≥ 10 mg/dL) (66%), lymphopenia (absolute lymphocyte count<1,000 per microliter) (50%), and hyponatremia (Na<135 mEq/L) (47%) – see Table 4.

**Table 1.** Demographic characteristics of convalescent plasma recipients^*^

|  | Overall, n=38 | Severe, 16 (42) | Critical, 22 (58) | p value |
| --- | --- | --- | --- | --- |
| Age, Mean (SD) | 63 (12) | 65 (11) | 61 (13) | 0.30 |
| Age Less than 70 | 28 (74) | 11 (69) | 17 (77) | 0.56 |
| Gender (female) | 18 (47) | 8 (50) | 10 (46) | 0.78 |
| Race |  |  |  | 0.92 |
| Black | 13 (34) | 6 (38) | 7 (32) |  |
| White | 12 (32) | 5 (31) | 7 (32) |  |
| Other | 13 (34) | 5 (31) | 8 (36) |  |
| Ethnicity (Hispanic) | 13 (34) | 4 (25) | 9 (41) | 0.31 |
| County |  |  |  | 0.45 |
| Hampden County | 15 (40) | 7 (44) | 8 (36) |  |
| Hartford County | 14 (37) | 7 (44) | 7 (32) |  |
| New Haven County | 8 (21) | 2 (12) | 6 (27) |  |
| Tolland County | 1 (3) | 0 (0) | 1 (5) |  |
| Insurance - Medicaid or Self-pay | 8 (21) | 4 (25) | 4 (18) | 0.19 |
| Marital Status - Married | 17 (45) | 7 (44) | 10 (45) |  |

\*Values are no. (%) except as indicated

**Table 2.**
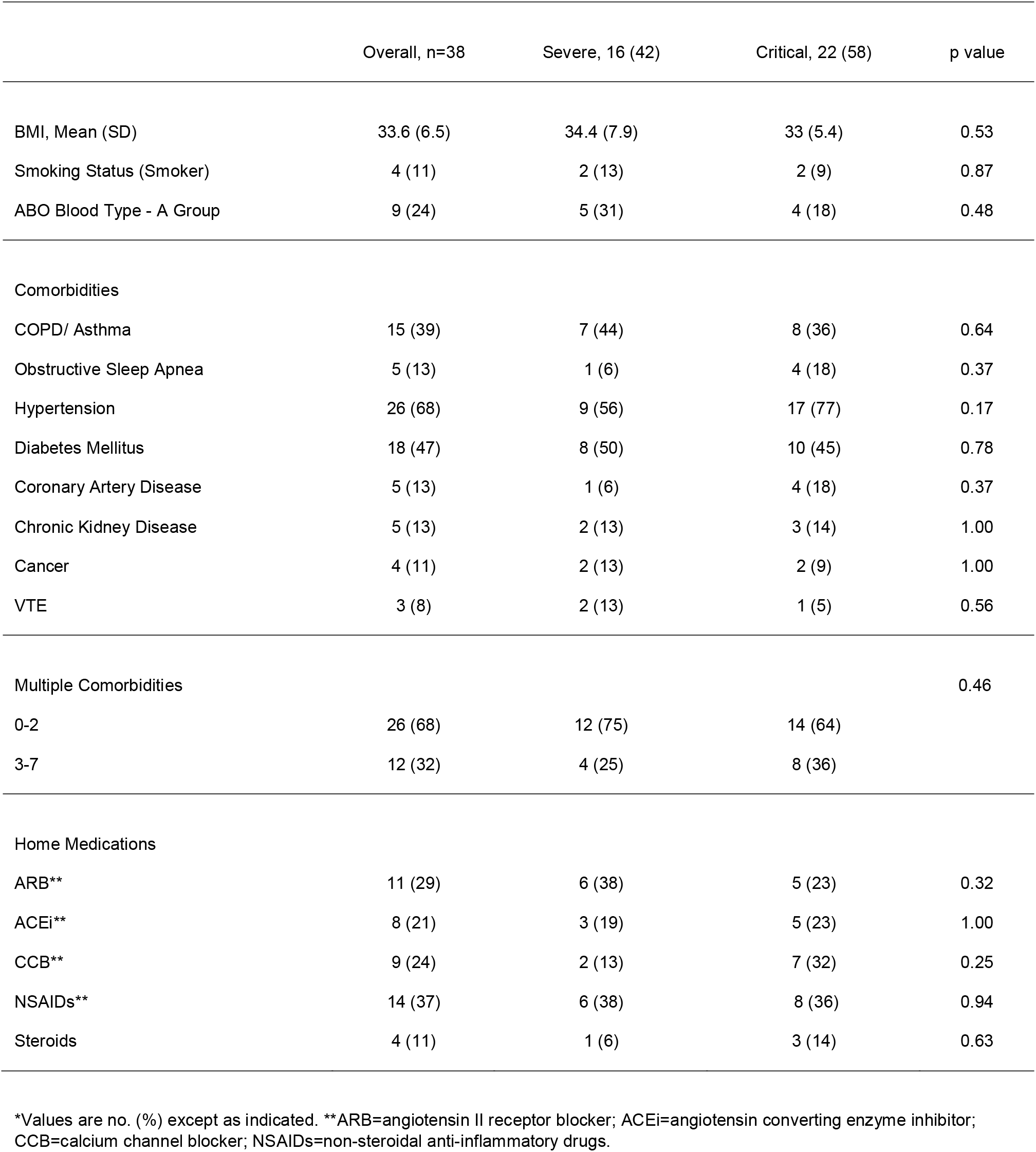
Comorbidities and home medications of convalescent plasma recipients^*^

**Table 3.**
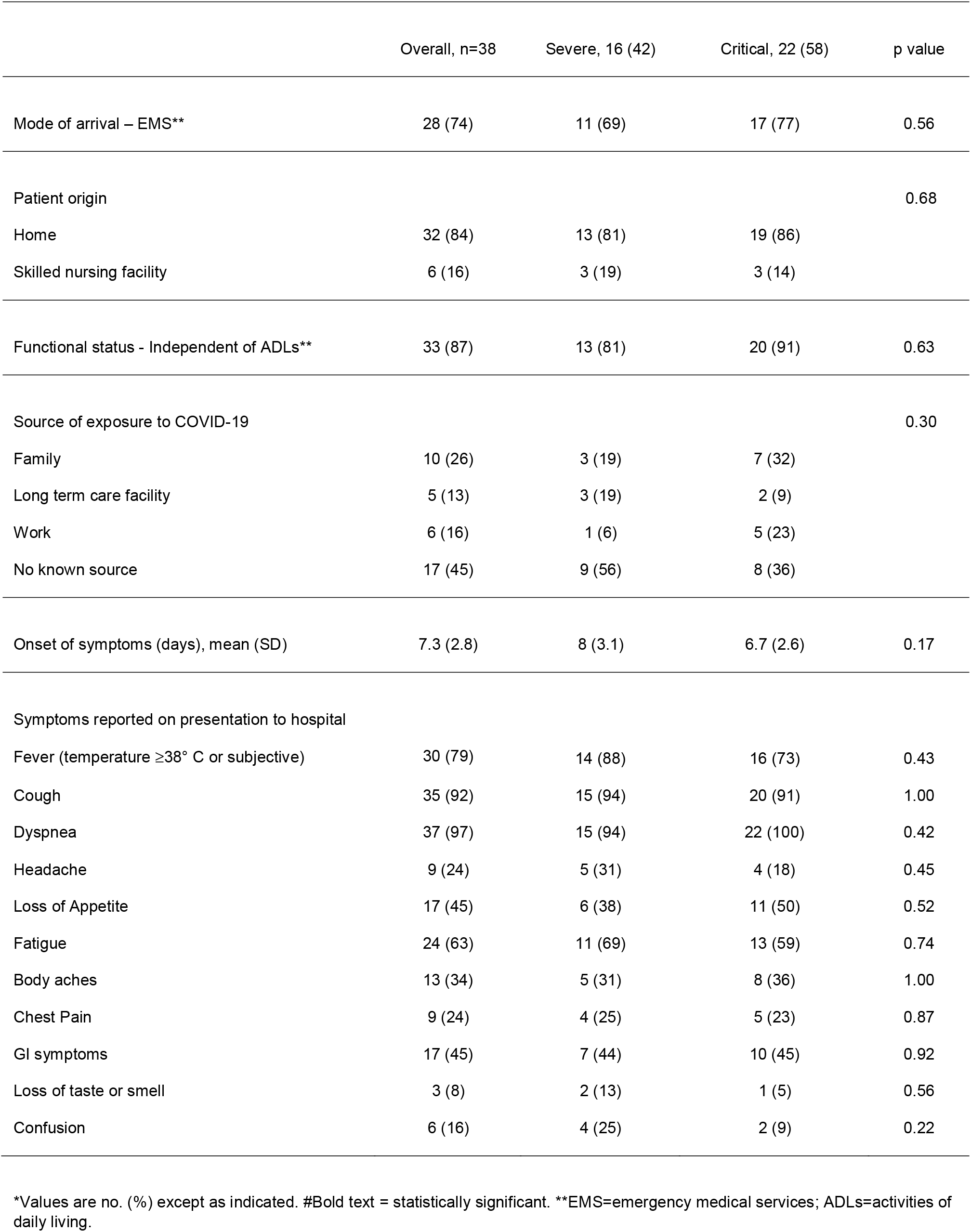
Characteristics and symptoms at presentation to the hospital among convalescent plasma recipients^*^

|  | Overall, n=38 | Severe, 16 (42) | Critical, 22 (58) | p value |
| --- | --- | --- | --- | --- |
| Mode of arrival – EMS** | 28 (74) | 11 (69) | 17 (77) | 0.56 |
| Patient origin |  |  |  | 0.68 |
| Home | 32 (84) | 13 (81) | 19 (86) |  |
| Skilled nursing facility | 6 (16) | 3 (19) | 3 (14) |  |
| Functional status - Independent of ADLs** | 33 (87) | 13 (81) | 20 (91) | 0.63 |
| Source of exposure to COVID-19 |  |  |  | 0.30 |
| Family | 10 (26) | 3 (19) | 7 (32) |  |
| Long term care facility | 5 (13) | 3 (19) | 2 (9) |  |
| Work | 6 (16) | 1 (6) | 5 (23) |  |
| No known source | 17 (45) | 9 (56) | 8 (36) |  |
| Onset of symptoms (days), mean (SD) | 7.3 (2.8) | 8 (3.1) | 6.7 (2.6) | 0.17 |
| Symptoms reported on presentation to hospital |  |  |  |  |
| Fever (temperature $\geq 38^{\circ}$ C or subjective) | 30 (79) | 14 (88) | 16 (73) | 0.43 |
| Cough | 35 (92) | 15 (94) | 20 (91) | 1.00 |
| Dyspnea | 37 (97) | 15 (94) | 22 (100) | 0.42 |
| Headache | 9 (24) | 5 (31) | 4 (18) | 0.45 |
| Loss of Appetite | 17 (45) | 6 (38) | 11 (50) | 0.52 |
| Fatigue | 24 (63) | 11 (69) | 13 (59) | 0.74 |
| Body aches | 13 (34) | 5 (31) | 8 (36) | 1.00 |
| Chest Pain | 9 (24) | 4 (25) | 5 (23) | 0.87 |
| GI symptoms | 17 (45) | 7 (44) | 10 (45) | 0.92 |
| Loss of taste or smell | 3 (8) | 2 (13) | 1 (5) | 0.56 |
| Confusion | 6 (16) | 4 (25) | 2 (9) | 0.22 |

\*Values are no. (%) except as indicated. #Bold text = statistically significant. \*\*EMS=emergency medical services; ADLs=activities of
daily living.

**Table 4.** Clinical presentation at admission of convalescent plasma recipients^*^

|  | Overall, n=38 | Severe, 16 (42) | Critical, 22 (58) | p value |
| --- | --- | --- | --- | --- |
| Disease severity (FDA Classification) on admission |  |  |  | 0.14 |
| Moderate | 5 | 4 (25) | 1 (5) |  |
| Severe | 32 | 12 (75) | 20 (90) |  |
| Critical | 1 | 0 (0) | 1 (5) |  |
| RA O2Sat ≤ 85%** | 18 (47) | 6 (38) | 12 (55) | 0.30 |
| Oxygen support on admission |  |  |  | 0.61 |
| Low Flow Nasal | 27 (71) | 11 (69) | 16 (72) |  |
| High Flow Nasal | 10 (26) | 5 (31) | 5 (23) |  |
| Invasive Ventilation | 1 (3) | 0 (0) | 1 (5) |  |
| Chest X-ray with typical COVID-19 findings** | 35 (92) | 14 (88) | 21 (95) | 0.56 |
| Febrile (temperature ≥38° C) | 10 (26) | 6 (38) | 4 (18) | 0.27 |
| Hypotension** | 1 (3) | 0 (0) | 1 (5) | 1.00 |
| Lymphopenia** | 19 (50) | 9 (56) | 10 (45) | 0.51 |
| Rise in sepsis markers** | 13 (34) | 4 (25) | 9 (41) | 0.31 |
| Severe rise in inflammatory markers** | 25 (66) | 10 (63) | 15 (68) | 0.71 |
| Transaminitis** | 9 (24) | 4 (25) | 5 (23) | 1.00 |
| AKI** | 10 (26) | 7 (44) | 3 (14) | 0.06 |
| Hyponatremia** | 18 (47) | 11 (69) | 7 (32) | **0.047** |
| Hypokalemia** | 7 (18) | 1 (6) | 6 (27) | 0.20 |
| Troponin leak** | 7 (18) | 2 (13) | 5 (23) | 0.68 |

\*Values are no. (%) except as indicated. #Bold text = statistically significant. \*\*RA O2Sat=room-air oxygen saturation; Chest X-ray with typical COVID-19 findings=multifocal peripheral consolidation and/ or multifocal founded opacities and nodules; Hypotension= mean arterial blood pressure less than 60 mm Hg; Lymphopenia=absolute lymphocyte count less than 1,000 per microliter; Rise is sepsis markers=serum lactate>2.2 mmol/L; Severe rise in inflammatory markers=C-reactive protein≥10 mg/dL; Transaminitis= 5x or greater rise in serum ALT level; AKI=acute kidney injury when eGFR<60 mL/min/1.73 sq meters; Hyponatremia=serum sodium less than 135 mEq/L; Hypokalemia=serum potassium less than 3.5 mEq/L; Troponin leak=serum troponin>0.04 ng/mL.

### Severe and Critical Illness Groups

At the time of plasma infusion, 16 patients (42%) met criteria for severe illness. These patients were enrolled in the study and received convalescent plasma earlier in their hospital and disease course on average 4.6 days (95% CI 2.9-6.3) following hospital admission, and 12.6 days (95% CI 10-15.2) following symptoms’ onset while on high-flow oxygen supplementation prior to any evidence of ARDS. The remaining 22 patients (58%) met the criteria for critical illness at the time of convalescent plasma therapy. They enrolled in the study and received convalescent plasma later in their hospital and disease course on average16.4 days (95% CI 13-19.8) following hospital admission, and 23.1 days (95% CI 19.5-26.7) following symptoms’ onset after developing ARDS and had been on ventilation support for an average of 10.6 days (95% CI 7.3-13.9).

The two cohorts were comparable in demographics; comorbidities and home medications; pre-illness functional status; onset of symptoms to seeking hospital care; initial clinical presentation and findings; initial disease severity; and the care they received during their hospitalization including essential medications (see Tables 1-5). Clinically, hyponatremia on initial hospital presentation was more prevalent in the severe illness group (p=0.047). Vasopressors (p<0.01), hydroxychloroquine (p<0.01), and antibiotics (p<0.01) were more frequently used during hospitalization in the critical illness group. Renal replacement therapy was utilized at higher rate in the critical illness group but did not reach statistical significance (p=0.05).

### Primary Outcomes

One patient in the severe illness group experienced a transient transfusion reaction (fever and hematuria) within 2 hours of plasma infusion. No other adverse effects of convalescent plasma infusion were observed. Of the 38 patients included in the analysis, 24 (63%) recovered and were discharged from the hospital, and 14 (37%) died. Patients who died included two in the severe illness group and 12 in the critical illness group. The difference in mortality (13% severe vs 55% critical) was statistically significant (p=0.02). Overall, patients who survived (n=24) regardless of disease severity at time of infusion received convalescent plasma earlier in their course of disease (mean 15.3 days, SD 6.9) and hospital stay (8.4 days, SD 6.8) compared to those who died (n=14) with mean durations of (24.5 days, SD 9.6), (16.6 days, SD 9.5) respectively.

### Secondary Outcomes

Among patients with severe illness at the time of convalescent plasma therapy, 25% (4/16) progressed to ARDS after receiving convalescent plasma (Table 5). Three of the 4 required mechanical ventilation and 2 of the 4 died. One of those patients received convalescent plasma 18 days following onset of symptoms and died of refractory shock in ICU while on ventilator support. The other patient received convalescent plasma 16 days following symptom onset, developed respiratory failure secondary to ARDS, and was placed on comfort measures at the request of the family. The remainder (14/16, 88%) did not progress to ARDS, recovered with resolution of COVID-19 pneumonia, and were discharged from the hospital.

**Table 5.** Patient Outcomes and Hospital Course^*^

|  | Overall, n=38 | Severe 16 (42) | Critical, 22 (58) | p value |
| --- | --- | --- | --- | --- |
| Outcome |  |  |  |  |
| Mortality | 14 (37) | 2 (13) | 12 (55) | **0.02** |
| Length of stay (days), mean (SD) | 26 (15) | 15.4 (11.6) | 33 (12.9) | **<0.01** |
| Symptom onset to CP (days), mean (SD) | 18.7 (9.0) | 12.6 (5.3) | 23.1 (8.6) | **<0.01** |
| Symptom onset to CP admin ≤ 15 days | 17 (45) | 13 (81) | 4 (18) | **<0.01** |
| Hospital days prior to CP admin, mean (SD) | 11.4 (8.8) | 4.6 (3.4) | 16.4 (8.1) | **<0.01** |
| Hospital days after CP admin, mean (SD) | 14.2 (11.5) | 10.9 (10.5) | 16.5 (11.9) | **<0.01** |
| ARDS** prior to CP admin | 22 (58) | 0 (0) | 22 (100) | **<0.01** |
| ARDS** during hospitalization | 26 (68) | 4 (25) | 22 (100) | **<0.01** |
| Invasive mechanical ventilation | 25 (66) | 3 (19) | 22 (100) | **<0.01** |
| Ventilator days, mean (SD) | 20.3 (10.3) | 16 (12.1) | 21 (10.2) | 0.45 |
| Other interventions and medications |  |  |  |  |
| Renal Replacement Therapy | 9 (24) | 1 (6) | 8 (36) | 0.05 |
| Antibiotics | 32 (84) | 10 (63) | 22 (100) | **<0.01** |
| Antifungals | 4 (11) | 0 (0) | 4 (18) | 0.12 |
| Azithromycin | 16 (42) | 4 (25) | 12 (55) | 0.07 |
| Hydroxychloroquine | 17 (45) | 3 (19) | 14 (64) | **<0.01** |
| IL-6 Inhibitors | 10 (26) | 3 (19) | 7 (32) | 0.47 |
| Remdesivir | 4 (11) | 3 (19) | 1 (5) | 0.29 |
| Vasopressors | 20 (53) | 3 (19) | 17 (77) | **<0.01** |
| Steroids | 22 (58) | 7 (44) | 15 (68) | 0.13 |
| Anticoagulants | 31 (82) | 13 (81) | 18 (82) | 1.00 |
| Zinc | 17 (45) | 6 (38) | 11 (50) | 0.44 |

\*Values are no. (%) except as indicated. #CP = convalescent plasma; \*\*ARDS = Acute respiratory distress syndrome; bold text =
statistically significant.

In the patients with critical illness at the time plasma therapy, 10/22 (45%) recovered with resolution of ARDS and restoration of organ function and left the hospital. Of the 12/22 (55%) who died, 6 died of refractory shock while on ventilator support with evidence of pneumoperitonium in 4 of them; 3 patients died of refractory respiratory failure with terminal extubation; 2 died of complications of upper airway edema; and 1 patient died of an acute cardiac complication.

Mean hospital length of stay was 25.6 days (95% CI 20.8-30.4) (Table 5). Length of stay was significantly shorter in the severe illness group (15.5 days, 95% CI 9.3-21.6) compared to patients in the critical illness group (33.0 days, 95% CI 27.3-38.7) (p<0.01). Statistical analyses showed that patients treated earlier in the course of COVID-19 disease (severe group) had significantly lower hospital mortality (p=0.02) and shorter hospital length of stay (p<0.01) after convalescent plasma therapy compared to patients that were treated later in their disease course in presence of ARDS (critical group) (Table 5). Other prognostic factors that were significantly associated with good clinical outcomes included shorter durations between symptoms onset and convalescent plasma administration (p<0.01), and hospital admission and administration of convalescent plasma (p<0.01).

## Discussion

Among this group of hospitalized patients with severe or critical COVID-19 who received convalescent plasma with adequate antibody titer, only one patient experienced a transient transfusion reaction. This low rate of adverse event secondary to convalescent plasma therapy is consistent with recent published literature (*22, 23*). The overall hospital mortality among our patients was 37%. However, patients who received convalescent plasma early in the disease course (severe illness group) as compared to the patients that received convalescent plasma later in disease progression (critical illness group) had significantly lower hospital mortality 13% vs 55% (p<0.02) and shorter mean hospital length of stay 15.4 vs 33 days (p<0.01). In addition, only 4 patients (25%) in the severe illness group developed ARDS, with 3 of them needing invasive ventilation support following convalescent plasma therapy. Two of the 3 recovered and were discharged.

It is important to understand the timeline and dynamics of COVID-19 hospitalizations in Connecticut and Western Massachusetts at the time when we launched our research study. Our study patients presented initially to the hospital with an average of 7.3 days (95% CI 6.4-8.2) from symptoms’ onset to hospitalization, and 97 % (37/38) of the patients had moderate to severe disease without evidence of ARDS or urgent need for invasive ventilation support upon admission. By the time we enrolled our first patient in late April 2020, hospitals participating in the study were at their peak COVID-19 census, with a large number of seriously ill patients who had been hospitalized for an average of 2 weeks, and were not improving with supportive care or medications (see Table 5). Some of those patients deteriorated and needed ICU care and ventilator support for an average of 7 days prior to enrollment. Many had severe lung damage and multi-organ failure. In the early phase of our study, physicians enrolled mostly patients in this critical illness category. In majority of cases, patients died due to secondary irreversible complications of COVID-19. As our study progressed, physicians started enrolling patients earlier in their disease course and hospital stay before respiratory status deterioration.

Upon admission to the hospital, the two cohorts were similar in their demographic characteristics, pre-illness functional status, comorbidities, initial clinical presentation to the hospital, and initial disease severity. However, at the time of convalescent plasma administration, the groups diverged on their disease severity and duration from disease onset to plasma therapy. The patients in our study who received convalescent plasma earlier in their disease course (severe illness group) had significantly more favorable primary and secondary clinical outcomes as compared to the critical group. We speculate that convalescent plasma given earlier in the disease course arrested the progression to irreversible complications like ARDS or organ failure. In addition, we found that patients who survived in both groups had shorter times between onset of symptoms and convalescent plasma administration compared to those that died.

The literature suggests that convalescent plasma may be more beneficial when administered sooner to disease onset (*16, 17*). Data recently published on COVID-19 suggested favorable clinical outcomes when convalescent plasma is given earlier in the course of disease (*21-25*). Our finding is consistent with the literature that treating patient with COVID-19 disease with convalescent plasma within the first two weeks following symptom onset may promote recovery (*27*). Perhaps earlier treatment with convalescent plasma allows antibodies to neutralize the virus before irreversible complications (*19*). Vasopressor, antibiotics and renal replacement therapy were utilized at higher rate in the critical group – we speculate that these therapies were proxies for serious and refractory complications among critically ill patients that could not be reversed by administration of convalescent plasma. We also speculate that the difference in hydroxychloroquine utilization between the two groups is likely a reflection of the change of evidence in association with hydroxychloroquine efficacy and safety and subsequent change in practice during our study. Important limitations in this study include open-label, no control group and modest sample size. However, this early report is important as it meaningfully contributes to the questions whether convalescent plasma is safe and it sheds light on important factors that are associated with favorable outcomes including recovery and survival.

### Conclusions

For patients with severe or critical COVID-19 disease, convalescent plasma from recovered COVID-19 patients is safe and has the potential for positive impact on clinical outcomes including recovery and survival if given early in course of disease. Our study makes a strong case for the importance of pursing a randomized placebo control trial focused on enrolling patients early in the course of their disease to further explore experimentally the efficacy and effectiveness of convalescent plasma in Covid-19.

## Data Availability

Data details is summarized and included in this submission. Patient level data without PHI is available upon request.

## Authorship

All named authors meet the International Committee of Medical Journal Editors (ICMJE) criteria for authorship for this article, take responsibility for the integrity of the work as a whole, and have given their approval for this version to be published.

We thank the donors, patients and their families for contributing to this work. We also thank the physicians from the Trinity Health Of New England Hospitals who enrolled patients in this study at Johnson Memorial Hospital, Stafford, Connecticut (Ian Tucker); Mercy Medical Center, Springfield Massachusetts (Robert Roose, Laurie Loiacono, and Vikram Sondi); Saint Francis Hospital and Medical Center, Hartford Connecticut (Phillip Roland, Jessica Abrantes-Figueiredo, Daniel Gerardi, Prashant Grover, and Gagandeep Singh), and St. Mary’s Hospital in Waterbury, Connecticut (Paul Porter and Bethel Shiferaw); the New York Blood Center and Rhode Island Blood Center for assisting with the plasma collection process; Ernst J. Schaeffer and Margaret R Diffenderfer, Boston Heart Diagnostics in Massachusetts, for serology testing; Collen Lima, Jessica McKenzie, Lisa Cook, Jennifer Puff, Mary Onoroski, and Patricia Nabors for their work with donors, specimen collection and plasma delivery; Kelly Batch, Donna Sobinski, Cynthia Considine, Brian Masthay, Christina Maxwell and Robert Wilke from the Trinity Health Of New England EPIC team for building the study in the electronic health record and assisting with study implementation. This study was funded and supported by Trinity Health Of New England, Hartford, Connecticut.

## Compliance with ethics guidelines

The study protocol was approved by the Trinity Health Of New England Institutional Review Board (#SFH-20-23). The research study was performed in accordance with the Helsinki Declaration of 1964, and its later amendments. Informed consent was provided by either the patient or the patient’s legally authorized representative (LAR).

## Data availability

The datasets generated during and/or analyzed during the current study are available from the corresponding author on reasonable request.

